# S-Press leg strengthening device in community inpatient wards: patient suitability, sit-to-stand ability, and patient-rated comfort outcomes

**DOI:** 10.1101/2024.12.24.24319402

**Authors:** Chris Griffiths, Kate Walker, Bharath Lakkappa

**Affiliations:** Research and Innovation Department, Northamptonshire Healthcare NHS Foundation Trust, Northampton, UK; Consultant/Clinical Director Community/CECS, Northamptonshire Healthcare NHS Foundation Trust, Northampton, UK

## Abstract

**Background:** Leg muscle deconditioning due to hospitalisation results in loss of muscle strength, physical immobility, and decreased ability to carry out activities of daily living. This causes delayed discharges, more nursing, carer, or social service support following discharge, possible discharge to care home rather than the patient’s own home, and increased risk of readmissions. Leg muscle deconditioning is associated with increased mortality, infections, and depression. Enhancing leg muscle strength should form part of in-patients’ rehabilitation plans. Progressive resistance training (PRE) offers a cost-effective way of preserving and improving muscle strength and function; however, it is not typically carried out in hospital.

**Objective:** To examine patient suitability, sit-to-stand ability, and patient-rated comfort outcomes of a leg strength training device (S-Press) in five community inpatient wards in England’s National Health Service (NHS).

**Methods:** Design: an open-label patient cohort design with no control group. Collection of the following data: reason for admission, number of times S-Press was used, number of repetitions, set up position used in, resistance levels used, increase in resistance level over use, time for five-times sit-to-stand, and patient reported comfort rating. Measures of sit-to-stand were conducted before use of S-Press and before discharge.

**Results:** An extensive range of admission reasons were recorded for 45 patients who used S-Press, indicating widespread suitability. Forty patients had information on set up position used in: 25 (62.5%) in chair, 13 (32.5%) on bed, and 2 (5%) in wheelchair. Out of 28 people who used S-Press more than once, the average number of S-Press sessions was 5.2, with 57% increasing resistance level and number of repetitions. Five-times sit-to-stand data collected for 16 participants showed all had a reduced length of time taken to sit-to-stand. There was an average reduction in five-times sit-to-stand of 17.4 seconds. Thirty-seven patients rated comfort out of 10, the most frequent rating was 10 out of 10 (15 out of the 37); average rating was 8.73.

**Conclusion:** The S-Press is beneficial for patients’ rehabilitation by offering PRE that is simple and easy to use and can be integrated as part of their rehabilitation plans. Patients with a wide variety of reasons for admission can use S-Press to strengthen their legs, either in a chair, wheelchair or bed as required. The majority of patients increased resistance and repetitions that they could do, indicating increased leg strength. Most patients find S-Press to be very comfortable to use. Five-times sit-to-stand improvements were seen for all those measured; speeding up this essential functional process is linked to increased mobility and ability to carry out activities of daily living. S-Press could be introduced to the most hospital wards.

## Introduction

Lack of physical mobility due to muscle deconditioning and suboptimal physical rehabilitation can cause delays in discharging patients from hospital (Hendy et al. 2012; Smith et al. 2020). Delays in discharge are costly to England’s National Health Service (NHS), which is estimated at £1.7 billion (Kings Fund, 2023). They are also associated with increased mortality, infections, and depression, and reductions in patients’ mobility and ability to engage in their activities of daily living upon discharge (Rojas-García et al. 2018). This highlights the importance of preventing muscle deconditioning due to hospitalisation and optimising physical rehabilitation.

Hospital-associated deconditioning (HAD) is where there are negative physical mobility consequences and loss of independence in activities of daily living because of hospitalisation (Smith et al. 2020). HAD can lead to increased risk of falls (Welch et al. 2024). There is high risk of HAD; prevention of HAD should be a priority but is often an unmet need (Smith et al. 2020; British Geriatrics Society, 2023). The muscle deconditioning process begins within two days, and there is a 2 to 5% decline in muscle mass per day when a patient does not walk, or their legs are inactive, with baseline strength reducing by up to 20% in one week (Brennan, 2024). The significant loss in skeletal muscle mass accelerates decline in physical mobility (Suetta et al. 2007). Sixty-eight percent of patients are discharged below their pre-admission level of physical functioning (Gill et al. 2009).

Physical inactivity is associated with medical complications and physical deterioration for individuals regardless of their age (Sáez de Asteasu et al. 2020). When individuals have no or low mobility during a period of hospitalisation, a decline in activities of daily living often occurs and there are increases in institutionalised care requirements and mortality rates (Brown et al. 2007). Deconditioned patients are three times more likely to be re-admitted within 30 days of their discharge, with de-conditioning associated with 47% of delayed discharges (Lim et al. 2006).

Muscle power training improves muscle strength and stimulates muscle hypertrophy (Cadore & Izquierdo, 2018). Resistance exercise is making muscles work against a weight or force (e.g., body weight, free weights, bands, exercise machines) to improve muscle strength (Chan et al. 2022). Progressive resistance exercise (PRE) is exercising muscles with a resistance exercise that is progressively increased as muscle strength improves (Liu & Latham, 2009). PRE has been found to increase muscle strength even for individuals who are particularly frail (Suetta et al. 2007). Older adults can achieve similar gains to younger individuals with PRE training, where substantial adaptive plasticity in skeletal muscle and the neuromuscular system has been demonstrated (Daly et al. 2017). PRE has been found to be safe, acceptable, and well tolerated in older post-acute inpatients who require rehabilitation (Coleman et al. 2021). Structured exercise programmes started early during hospitalisation can prevent muscle strength and function deterioration, reversing the progressive impairment of patients’ functional ability for activities of daily living during a hospital stay, and can have benefits on cognitive functioning and quality of life (Martínez-Velilla et al. 2019).

There is substantial evidence for the benefits of using PRE (Liu & Latham, 2009), yet physiotherapists working in hospitals may not be routinely using it (Lim et al. 2006; Suetta et al. 2007). Barriers can be institutional-related, lack of physiotherapists, lack of appropriate equipment, and time pressures (So & Pierluissi, 2012; Parker et al. 2013; ShahAli et al. 2023). Other barriers include perceptions by staff of poor patient capacity to use PRE and a lack of confidence in using PRE as an intervention (Williams & Denehy, 2019), a lack of prioritisation of resistance exercises, perceived poor patient motivation, the belief by staff that some patients were unable to perform resistance exercises, a lack of definition around resistant exercises, and insufficient support staff for effective implementation (Chan et al. 2022). There are also barriers that exist to providing physiotherapy generally as part of in-patient rehabilitation such as patient fatigue; patient dependency; patients feeling weak, unwell, unstable, and/or anxious; pain; and pressure sores (Chan et al. 2022). Patient barriers causing low adherence with their exercise plans include low self-efficacy, depression, anxiety, and increased pain levels during exercise (Jack et al. 2010).

Barriers need to be reduced to enable PRE to be used more in hospitals to prevent deconditioning and improve muscle strength, and in doing so improve rehabilitation outcomes (Falvey et al. 2015). Preserving muscle strength and function should form part of patients’ rehabilitation plans, but there is a lack of PRE equipment available for physiotherapists. To seek to address this, a portable leg strengthening device called the Strength-Press (S-Press) has been designed by a physiotherapist, with the aim of improving access to and availability of effective PRE for adults in hospital. The device was designed to be easy to use from a professional and patient perspective, the S-Press can be used independently by appropriate patients once they are set up (Maden-Wilkinson et al. 2024). The S-Press is designed to remove barriers to effective rehabilitation, it is portable, safe and easy to set up, simple to use, and can be used by patients who are weak, frail, anxious and in pain (Maden-Wilkinson et al. 2024). The aim of this study was to examine patient suitability, sit-to-stand outcomes, and patient-rated comfort outcomes of the S-Press in five community inpatient wards in the NHS.

## Methods

### Design

Open-label patient cohort design with no control group.

### Approval

The project was undertaken from October 2023 to November 2024. Approval for the study was gained from the United Kingdom (UK) NHS healthcare trust in which the services were based: Northamptonshire Healthcare NHS Foundation Trust (NHFT). The NHFT ‘Innovation Forum’ committee approved (reference number: IF:S-Press). All participants provided informed consent. The study was delivered in accordance with the declaration of Helsinki.

### Inclusion and exclusion criteria

Inclusion criteria for the study were any NHS in-patient admitted to a ward site, assessed as medically stable and physically able to participate, and able to provide informed written consent for participation. Exclusion criteria included any patient unable to give informed consent, significantly confused patients, those unable to do leg press exercises, anyone acutely unwell with an infection, or with high blood pressure that is uncontrolled.

### Procedure

Potential participants were offered S-Press based in inclusion/exclusion criteria. Participants read the information sheets and considered the information prior to consenting to participate in the study. Collection of the following data by hospital staff was carried out: reason for admission, number of times used, number of repetitions, set up position used in, resistance levels used, increase in resistance level over use, time for five-times sit-to-stand, and patient-reported comfort rating.

### Setting: Adult Community Inpatient Wards

Five community hospitals offering inpatient specialist services for adult patients who need 24-hour inpatient care and inpatient levels of treatment, physical therapy, and support. The services are for sub-acute medicine and attend to a combination of medical, physical, and neurological needs. The patients may have had a severe illness, a decline in their health after a long-term illness, a stroke, or a brain injury, or need medical, physical or specialist rehabilitation. To meet these varied needs, ward teams are made up of a wide range of healthcare professionals. Some professionals are based at specific hospital sites and others come to hospital wards depending on a specific patient’s needs. Typically, there are twelve beds per ward.

### Training

JT Rehab physiotherapist attended each ward to do a 30-minute training session with staff who would be using the S-Press This training involved going through the S-Press protocol (as shown below), which includes how to set up and use the S-Press with patients. Those not on shift or unable to attend were taught by a colleague who attended the training. A training log was kept updated of those who underwent the training and who were competent to use the S-Press with patients.

### S-Press protocol

1. Review inclusion and exclusion criteria to see if patient meets the criteria set.
2. First time used: provide information sheet to patient, discuss and answers questions and gain written consent from patient.
3. Clean the S-Press prior to each use.
4. Carry S-Press to patient.
5. If patient is in bed – use bed set up information.
6. If patient is sitting in the chair – use chair set up information.
7. Assess the maximum resistance that the patient can achieve (1 repetition max).
8. Reduce resistance level to one below assessed 1 repetition max.
9. Instruct the patient to do 10 controlled leg extensions and returning to start line with knee flexed, on each leg firstly for the knee extensors and then (if in patient’s rehabilitation plan) the knee flexors.
10. National guidance for resistance training is recommended to be at least twice a week. Consenting patients can use the S-Press two or three times a week for as many weeks as they are an in-patient on the ward.

### Sit-to-stand

Sit-to-stand five times were calculated before the first S-Press session and prior to the patient’s discharge. Instructions given to the patient comprised: 1) Sit in the middle of the chair; 2) Place your hands on the opposite shoulder crossed, at the wrists; 3) Keep your feet flat on the floor; 4) Keep your back straight and keep your arms against your chest; and 5) On “Go,” rise to a full standing position, then sit back down again.

### Resistance levels used

Staff made a written record of set up position used in, number of times used, number of repetitions for each session, and resistance levels used for each session.

### S-Press Device

The S-Press is a Class 1 medical PRE device to strengthen the leg muscles, designed by a physiotherapist. The S-Press provides 3 - 29kg of therapeutic level resistance across six changeable levels. The S-Press works bidirectionally, targeting the quadriceps with use in one direction, and by rotating the device targets the hamstrings. The S-Press can be used on the bed, in a chair or wheelchair.

## Results

### Participants

Forty-five participants in total, 22 (63%) males and 13 females (37%), were recruited. The age range of participants was 32 to 94 years old, with a mean age of 74. Table 1 lists reported admission reasons.

**Table 1.** Reported admission reasons.

| Admission reason | Number of patients |
| --- | --- |
| Chronic inflammatory demyelinating polyradiculoneuropathy (CIDP). Falls | 1 |
| Pelvic, L4 /L5 spinal, Punctured bladder, L sacral plexopathy | 1 |
| Asthma exacerbation, Functional Neurological Dysfunction (FND) | 1 |
| Leg ulcers and vasculitis | 1 |
| L5/S1 disc prolapse, severe low back pain | 1 |
| Knee Osteoarthritis | 1 |
| Neck of femur R – hemiarthroplasty | 1 |
| Fall. Neck of femur fracture. Dynamic hip screw fixation | 2 |
| Fall. Neck of femur fracture. | 3 |
| Fall. Ankle pain and swelling | 1 |
| Falls. | 2 |
| Fall. Acetabular | 1 |
| Fall. Decreased mobility and balance | 1 |
| Cardiovascular problems | 1 |
| Fall. Neck of femur. Hemiarthroplasty | 1 |
| Stroke | 10 |
| Fall. Subdural haematoma | 1 |
| Dense right hemiplegia | 1 |
| Fall. Distal humerus. Shingles | 1 |
| Oesophagitis | 1 |
| Infected leg ulcers | 1 |
| Peri-prosthetic. Chronic leg ulcers | 1 |
| Community-acquired pneumonia (CAP) and Non–ST-Segment Elevation Myocardial Infarction (NSTEMI) and Acute kidney injury (AKI) | 1 |
| Constipation and abdominal pain | 1 |
| Total Knee replacement revision surgery | 1 |
| Leg cellulitis and group B strep. Knee pain. Acute kidney injury (AKI) | 1 |
| Clostridioides difficile (C.diff). Diarrhoea | 1 |
| Low back pain. Decreased mobility | 1 |
| Infected ulcer | 1 |
| Chronic inflammatory demyelinating polyneuropathy | 1 |
| Parkinson's. Reduced mobility and diarrhoea | 1 |
| Total knee replacement revision surgery | 1 |

## Results

An extensive range of admission reasons were recorded. Stroke was the most common admission reason (10 participants), followed by Falls and Neck of femur fractures. Out of 28 people who used the S-Press more than once, 57% increased resistance level and number of repetitions over their sessions. The average number of S-Press sessions for the 28 participants was 5.2.

Five-times sit-to-stand data showed all (100%) (*n*=16) we had data on reduced length of time taken to sit-to-stand, and one person was able to walk with the aid of a walking frame after using the device when previously unable to. There was an average reduction in five-times sit-to-stand of 17.4 seconds.

Thirty-seven patients rated comfort out of 10, the most frequent rating was 10 out of 10 (15 out of the 37); average rating was 8.73. Forty patients had information on set up position used in: 25 (62.5%) in chair, 13 (32.5%) on bed and 2 (5%) in wheelchair.

## Discussion

This study found that the S-Press is beneficial for patients’ rehabilitation by offering PRE that can be integrated as part of patients’ rehabilitation plans. Patients with a wide variety of reasons for admission can use the S-Press to strengthen their legs, either in a chair, wheelchair or bed as required, indicating wide ranging suitability. The majority of patients increased resistance and repetitions over their sessions, and therefore increased their leg strength. Most patients found the S-Press to be comfortable to use. Five-times sit-to-stand improvements were seen for all those reporting, better enabling this essential functional process of increased mobility. The findings support previous studies in post-acute inpatient populations showing improvements in muscle strength and function following bed-based (Mallery et al. 2003; Kawakami et al. 2001) and chair-based PRE (Latham et al. 2003; Suetta et al. 2007; Coleman et al. 2021).

Interviews with patients, carers and staff using S-Press have shown that the S-Press offers an accessible, easy-to-use form of PRE that can be routinely used in hospital (Walker et al. 2025). The S-Press was found to be portable and easy to manoeuvre, adaptable for use as per the patients’ needs (in bed, sitting) and could be used in place of other pieces of equipment that were less accessible (e.g., motorised cycle devices, gym base equipment) (Walker et al. 2025). Most patients reported that they were motivated to use the S-Press and were able to do the exercises on it; the S-Press provided exercise goals for them to work towards (Walker et al. 2025). The results from this study add to these qualitative findings, demonstrating improvements in indicators of leg strength.

Goal-setting is integral to rehabilitation interventions (Gayton & Monga, 2023), and a key tenet of patient centred musculoskeletal physiotherapy (Hutting et al. 2022). The S-Press can provide patients an objective measure of progress and feedback, increasing motivation and self-efficacy, as well as giving them hope (Walker et al. 2025). The S-Press was reported by physiotherapists to be a valuable contribution to strengthening patients’ leg muscles and as a result helped improved stability, balance, standing, and walking (Walker et al. 2025).

The combination of previous qualitative and these quantitative results support the use of the S-Press. Recent NICE guidelines (NICE, 2023) have recommended that needs-based rehabilitation for people after a stroke should be at least three hours a day, five days a week, covering a range of multidisciplinary therapy; the S-Press can be routinely used and contribute towards achieving this. Such guidance for other conditions would support the systematic use of PRM during in-patient settings.

### Limitations

There was no control group, a relatively small sample size, and the use of S-Press was open-label and adjunct to other treatments or therapies. This study collected outcome measures during hospitalisation only. The participants were from a single English county reducing generalisability. There was an under representation of females and so the results are less generalisable to females.

### Future research

Future studies could build on the qualitative and quantitative evidence for S-Press by including a validated measure of functioning or activities of daily living. A fully powered multi-site randomised controlled trial (RCT), comparing the results of a group who use S-Press only and a control group who received treatment as usual or another exercise device would provide additional evidence for S-Press effectiveness. Furthermore, a longer follow-up time would help to find out whether any improvements to leg strength, physical mobility and activities of daily living are sustained. Evaluation of the use of S-Press in the community, pre and post hospital admission would be valuable.

## Conclusion

The S-Press could be used in most in-patient wards to help prevent hospital associated deconditioning, strengthen legs, reduce risk of falls, enable and speed-up rehabilitation, enable more effective physiotherapy, reduce length of stay, enable discharge with less support than would be required without it, and enable discharge to own home rather than supported living. The use of the S-Press links to new UK governments’ focus on ‘making better use of technology in health and care’, ‘preventing sickness, not just treating it’ and ‘NICE guidance aligned integrating physical activity in clinical pathways’ (Department of Health & Social Care, 2024).

## Data Availability

All data produced in the present study are available upon reasonable request to the authors

